# Using Hourly Aggregated Respiratory Rate and Expiratory Time with Machine Learning to Identify Remote COPD Exacerbations

**DOI:** 10.1101/2025.10.24.25338693

**Authors:** R. Smunyahirun, T. Filhol, T. Trakoolwilaiwan, R. Al-Ani, M. Lynn, L. Liang, G. Balasundaram, J. Lombay, G. Singh, M. O. Wiens, P. Moschovis, M. Olivo

## Abstract

Exacerbations of chronic obstructive pulmonary disease (COPD) are a major cause of morbidity and mortality. Various models for identifying exacerbations have been proposed, but few models are based on continuous and passively recorded respiratory physiology data. We enrolled 17 subjects with COPD with at least 2 exacerbations in the prior 12 months. We developed machine learning models using aggregated respiratory rates and expiratory times collected passively in a home-setting to identify COPD exacerbations. Models achieved areas under the ROC curve of 0.7989 and 0.8720 respectively, for exacerbations defined by self-reported outcomes and COPD Assessment Test (CAT) score changes. This pilot study demonstrates the feasibility of using passively recorded respiratory physiological data to identify COPD exacerbations.

## 1. Introduction

Chronic obstructive pulmonary disease (COPD) is a long-term chronic respiratory disease which is a leading cause of healthcare utilization and mortality in the US, primarily driven by acute exacerbation events [1], [2]. Recurrent acute exacerbations of COPD (AECOPD) can lead to hospital readmission, accelerated lung function decline, and an increased risk of death [3], [4]. Therefore, the identification of AECOPD events as early as possible before symptom onset carries significant potential benefit, allowing for early outpatient intervention to avert potential additional healthcare utilization and further decline in lung function.

Various methods have been proposed for identifying AECOPD events. Patient-reported outcome measures (PROMs) [5], [6] such as the COPD Assessment Test (CAT), are associated with lung function decline and recurrent hospitalizations [7]. However, PROMs are also less likely to be consistently reported over longer periods of monitoring (e.g. 60 days [8], [9]). Pulse rate and oxygen saturation have also been used to identify exacerbations, with performances with area under the ROC curve (AUC) of 0.578 and 0.658 for pulse rate and oxygen saturation, respectively [10]. For oxygen saturation specifically, the change in oxygen saturation associated with an exacerbation can be 1-2% [11], which is within the range of error of pulse oximeters. Pulmonary function tests (PFT) such as spirometry have also been used to monitor peri-exacerbation lung function decline [12], but require active participation by the user, are effort- and technique-dependent [13], and several studies have demonstrated poor long-term adherence to home spirometry [14]. Furthermore, its performance in home settings, including validity, reliability, and interpretability, requires further evaluation to ensure accuracy compared to in-laboratory standards [15]. Daily respiratory rate measurement has also been shown to be able to identify home COPD exacerbations [16], though a single measurement lacks the ability to capture fluctuations within the day, such as differences between morning and night.

In this paper, we present a novel method for identifying COPD exacerbations by aggregating hourly respiratory rates and expiratory times at multiple timepoints within the day using machine-learning algorithms. The novelty of the method derives from use of respiratory rates and expiratory times at multiple timepoints within the day, including them as features in machine-learning algorithms to identify COPD exacerbations. The usefulness of the algorithm is that (1) it can potentially be used in conjunction with a passive wearable device to identify exacerbations rather than a solution that requires active engagement by the patient (e.g. PROMs through mobile applications/PFTs), and (2) it can perform better in a cohort of respiratory patients compared to wearables that measure non-respiratory parameters, e.g., pulse oximetry [17].

## 2. Methods

### 2.1 Study Population

We enrolled 17 subjects diagnosed with COPD at the Massachusetts General Hospital (MGH) and Salem Hospital in a prospective cohort/diagnostic accuracy study approved under the Mass General Brigham Institutional Review Board (IRB Protocol #2021P000895, Clinicaltrials.gov ID: NCT04825067). Inclusion criteria were at least 2 documented exacerbations in the last 2 months. Exacerbations were defined following the Global Initiative for Chronic Obstructive Lung Disease (GOLD) guidelines, as increase in frequency and severity of cough, increase in volume and/or change of character of sputum production, or increase in dyspnea; and treatment with short-acting bronchodilator, antibiotics, and oral or intravenous glucocorticoids [18]. We excluded potential subjects with a history of adhesive or tape allergy, neuromuscular diseases, subjects enrolled in hospice care and subjects who lived more than 60 miles away from the enrollment site. All subjects provided written informed consent.

### 2.2 Study procedures

A U.S. Food and Drug Administration (FDA)-approved chest patch-based wearable device (RS001 Cardio-respiratory Monitor, Respiree, Singapore) was used to measure respiratory rate and expiratory time in a continuous manner. The wearable device is an optical sensor based on a direct-contact optical diffuse reflectance phenomenon [19]. Subjects were trained on use of the device prior to initiating the study.

Subjects were asked to wear the Respiree device on the upper chest for between 30 and 90 days. Subjects were asked to charge the device twice daily, once in the morning and once at night before attaching the device prior to sleep. A mobile phone running a gateway application was also provided to subjects to automatically synchronize physiologic data from the device to the phone via Bluetooth, and data was then uploaded to the cloud-based database. Physiologic waveform data were then processed to provide respiratory rates and expiratory times at resolutions of two minutes.

At baseline and on a weekly basis, subjects completed questionnaires which included CAT scores, any worsening of symptoms, healthcare utilization, and changes in medications. Questionnaires were administered either via a web-based interface (REDCap [20], [21]) or via phone interview for those who preferred to do so. In this study, we considered 2 types of outcomes as follows: (1) a change in weekly CAT score by +4 and (2) self-reported worsening of symptoms on a weekly basis. A change in weekly CAT score by +4 points was used, which was twice the magnitude of the reported minimum clinically important difference of +2 [22].

### 2.3 Development of the Algorithm

The algorithm was developed to identify pre-exacerbation days in COPD patients through remote monitoring [23]. The algorithm consists of two components: data pre-processing and the machine learning model. The machine learning models were trained and tested on datasets independently.

#### 2.3.1 Data Pre-processing

Respiratory rate and expiratory time were captured every two minutes from the RS001 monitor, along with signal quality and on-skin status. Signal quality was categorized into four levels (good, moderate, poor, and motion) based on raw accelerometer data captured by the sensor [19]. The sensor also provided a Boolean value indicating the on-skin status, determining whether the sensor was worn and in contact with the skin. To filter out noise, only respiratory rate and expiratory time with good or moderate quality and on-skin status were used for model development.

Filtered data points were then averaged over three-hour windows to reduce noise due to minute-level fluctuations in respiratory rate and expiratory time.

To concatenate the predictor across multiple time points, we introduced an observation window. Within the observation window, all aggregated data were concatenated into a feature vector. The length of the feature vector (i.e., the number of features), depends on the window size. For example, an observation window of five days consists of 40 features in total, as each day has eight points of three-hour aggregated data.

,where *X* is the feature matrix, *n* the five-hour observation window number, and *RR* the average Respiratory Rate over a three-hour window.

The observation window slides from the beginning to the end of the data with a one-day stride, creating multiple feature vectors for each subject (c.f., Figure 1 and Figure 2)

**Figure 1.**
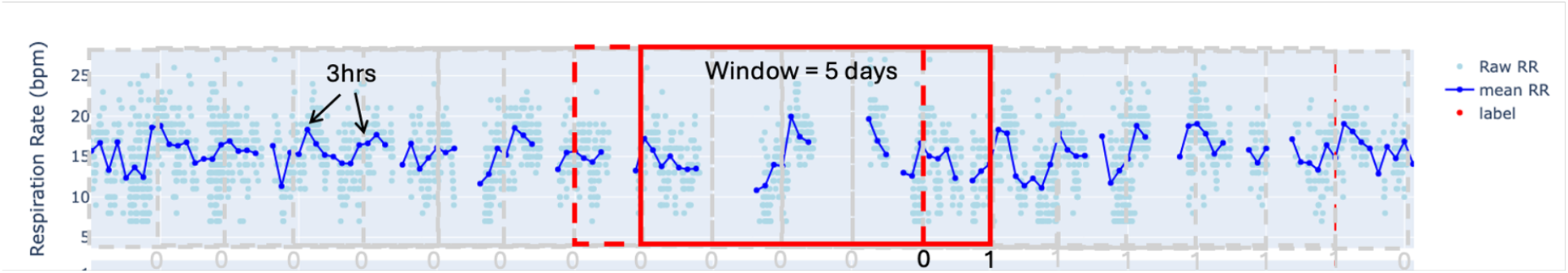
Observation window of 5 days

**Figure 2.**
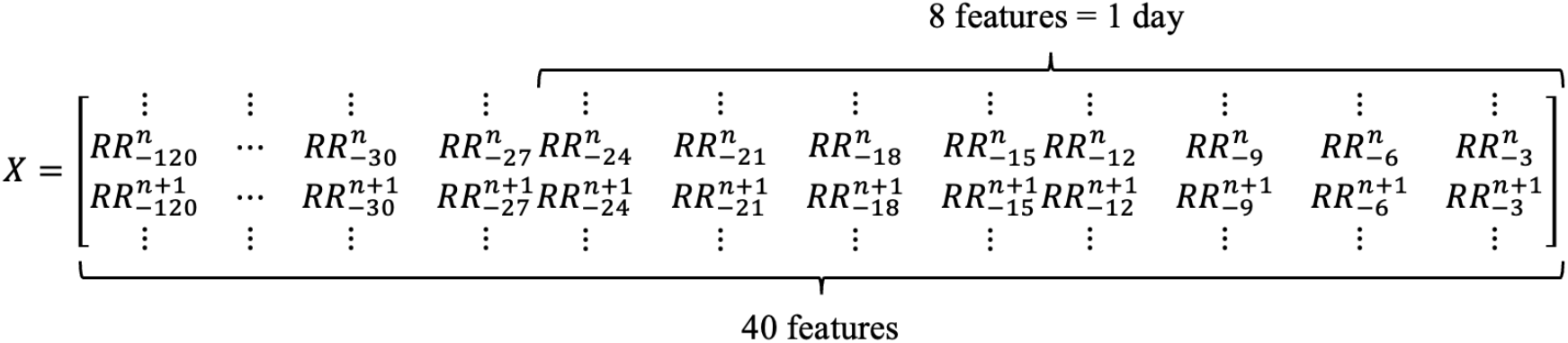
Breakdown of the 40-feature vector

To minimize missing data points in a feature vector, we calculated the minimum data threshold expressed as a percentage. For example, a minimum data threshold of 25% for a five-day window means that at least 10 aggregated data points (25% of 40 data points) need to be present in the window; otherwise, the feature vector corresponding to the window is discarded.

#### 2.3.2 Data Labeling

We labeled the five days preceding an exacerbation event as well as the day of the event itself as exacerbation days. These labels at the prediction time of the observation window served as the ground truth for the corresponding feature vector. In machine learning terminology, feature vectors labeled as exacerbation days are referred to as positive vector samples, while those that are not, are called negative vector samples. Figure 3 illustrates the data pre-processing and the data labeling.

**Figure 3.**
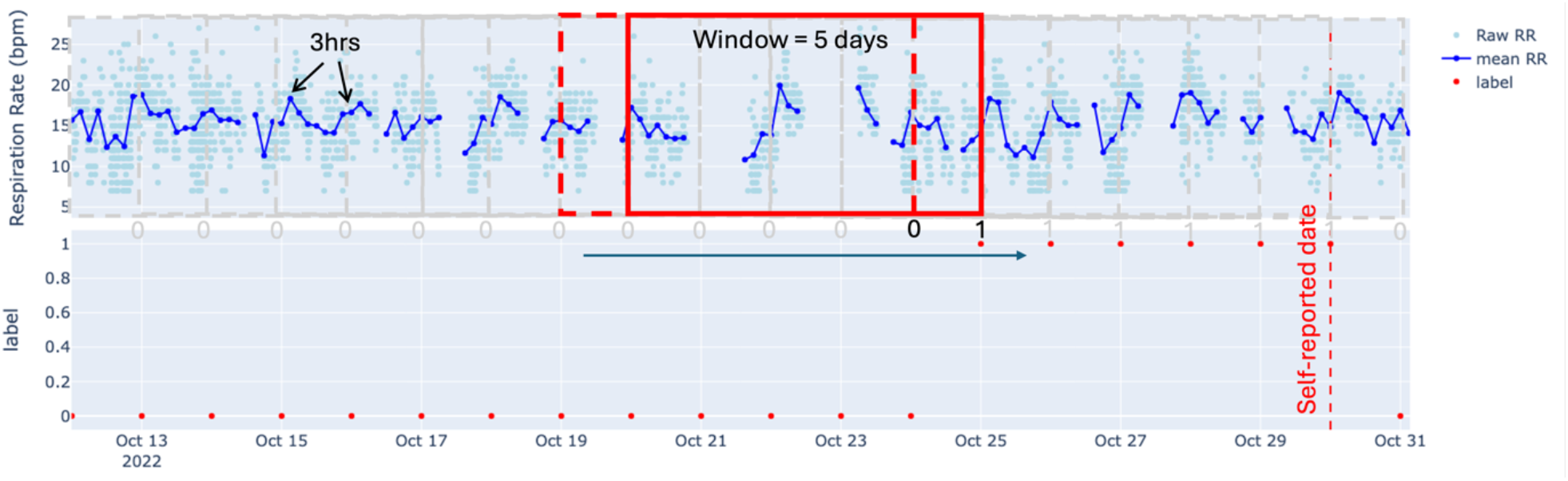
Observation window of 5 days with labels, indicating whether a patient will experience exacerbation the following day.

#### 2.3.3 Train-Test Split

The vector samples were split into an 80% training set and a 20% test set. Due to the imbalanced nature of the data, a stratified split was applied. The data was split in two ways: fold split and patient split.

The fold split is a conventional method where each vector sample is treated independently. With a 20% test set, this method provided five folds. However, since a single subject generated multiple vector samples, the fold split could result in vector samples from the same patient being included in both the training and test sets, potentially causing data leakage into the test set.

To prevent data leakage, a patient split was also implemented. This can also be defined as a patient-level fold. This method splits the vector samples by patient, ensuring no data leak into the test set while maintaining a stratified split by constraining the difference in the ratio of positive vector samples between the training and test sets to be less than 1%. Given this constraint, there were limited combinations of training and test sets available.

Additionally, the number of positive vector samples and the total number of vector samples depend on data cutting parameters which are the observation window and the minimum data threshold, as described in the previous section. Different parameter values could lead to different combinations of train-test sets.

Tables A1 and A2 in the supplementary data provide detailed descriptions of the train-test sets for each splitting method for self-reported outcome and CAT score changes outcome with optimal data cutting parameters which are observation window and minimum data threshold. Note that the observation window size is independent of the labeling strategy where five days preceding exacerbation events are the pre-exacerbation days. Details of optimization will be explained in a later section. For the self-reported outcome, there are five train-test sets with an observation window of five days and a minimum data threshold of 50%. For the CAT score changes outcome, there are twelve train-test sets with an observation window of four days and a minimum data threshold of 25%.

#### 2.3.4 Model Training

Extreme Gradient Boosting, commonly known as XGBoost, is one of the most popular and robust machine learning models for binary classification tasks. It is an ensemble tree-based learning method. We trained the XGBoost models and performed hyperparameter tuning for each train-test set independently. The hyperparameters we optimized included learning_rate, scale_pos_weight (the weight of positive samples), and max_depth, while other parameters were kept at their default values as per XGBoost package version 2.0.3. The values of the hyperparameters used in the grid search are shown in Table 1. The grid search was executed to identify the set of hyperparameters that maximized the average precision score of the test set.

**Table 1.**
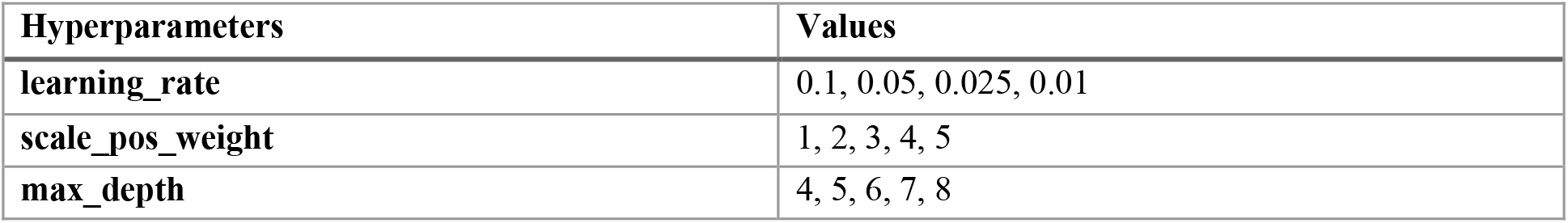
Values of Parameters for Hyperparameters Tuning of XGBoost.

| Hyperparameters | Values |
| --- | --- |
| **learning_rate** | 0.1, 0.05, 0.025, 0.01 |
| **scale_pos_weight** | 1, 2, 3, 4, 5 |
| **max_depth** | 4, 5, 6, 7, 8 |

For comparison, we also trained a logistic regression model on the patient split. Hyperparameter tuning was conducted for this model as well, with the values shown in Table 2 while the other parameters were kept at their default values as per Skicit-learn package version 1.4.1.post1. However, since the logistic regression model cannot handle missing values, all missing values were imputed using the average of the predictor values within the observation window.

**Table 2.** Values of Parameters for Hyperparameters Tuning of Logistic Regression.

| Hyperparameters | Values |
| --- | --- |
| **C** | 0.0001, 0.001, 0.01, 0.1, 1.0 |
| **l1_ratio** | 0.0, 0.1, 0.2, 0.3, 0.4, 0.5, 0.6, 0.7, 0.8, 0.9, 1.0 |
| **class_weight** | 2, 3, 4, 5, 6 |

Additionally, we performed a grid search not only on the model hyperparameters but also on the data cutting parameters, namely the observation window size and the minimum data threshold. The values for window size and minimum data threshold are shown in Table 3.

**Table 3.**
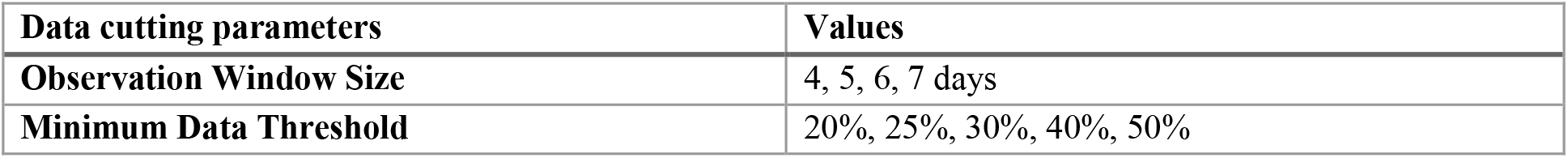
Values of Data Cutting Parameters for Hyperparameters Tuning.

| Data cutting parameters | Values |
| --- | --- |
| **Observation Window Size** | 4, 5, 6, 7 days |
| **Minimum Data Threshold** | 20%, 25%, 30%, 40%, 50% |

It is possible for aggregated data to be missing, indicating that no data was captured for the entire three-hour period. XGBoost can handle missing data naturally. Therefore, we retained the missing data without any imputation.

#### 2.3.5 Model Evaluation

The model was evaluated using the following set of metrics: ROC AUC, accuracy, recall, precision, specificity, and F1 score [24].

ROC AUC, which stands for Receiver Operating Characteristic - Area Under the Curve, assesses a model’s ability to distinguish between two classes (e.g., positive or negative) across various threshold settings. The ROC curve is the plot of the true positive rate (TPR or Recall) against the false positive rate (FPR) at each threshold setting (Equation (E1), (E2) in the supplementary data).

The accuracy score is a metric that represents the proportion of correct predictions a model makes out of the total number of predictions (Equation (E3) in the supplementary data).

Recall is a metric that measures how well a model identifies positive instances from all the actual positive samples in a dataset (Equation (E4) in the supplementary data).

Precision is a metric that measures the accuracy of positive predictions made by a model (Equation (E5) in the supplementary data).

Specificity indicates the proportion of actual negative cases that are correctly classified as negative (Equation (E6) in the supplementary data).

The F1 score represents the harmonic mean of precision and recall, providing a balanced measure that considers both false positives and false negatives (Equation (E7) in the supplementary data).

## 3. Results

### 3.1 Dataset

The demographic of 17 enrolled subjects is shown in Table 4. To identify days leading up to an exacerbation, we labelled a maximum of 5 days leading up to an exacerbation as exacerbation days. The total number of exacerbation events, days, and percentage of exacerbation days of the total possible days is shown in Table 5. If 2 exacerbations occurred within the labelled number of days (assuming it is 5), then the labelled number of days was added between the 2 exacerbation events.

**Table 4.**
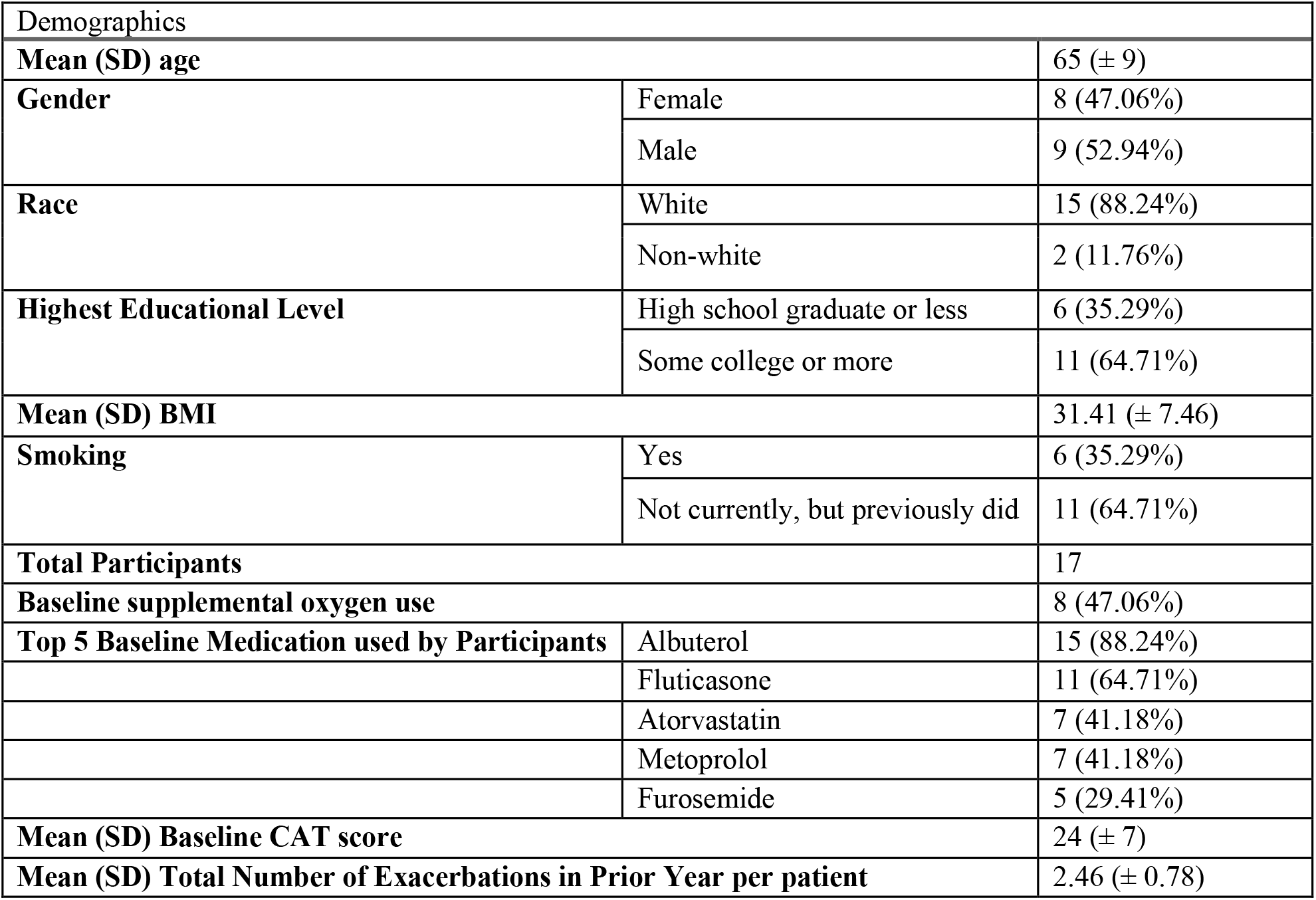
Demographics.

**Table 5.**
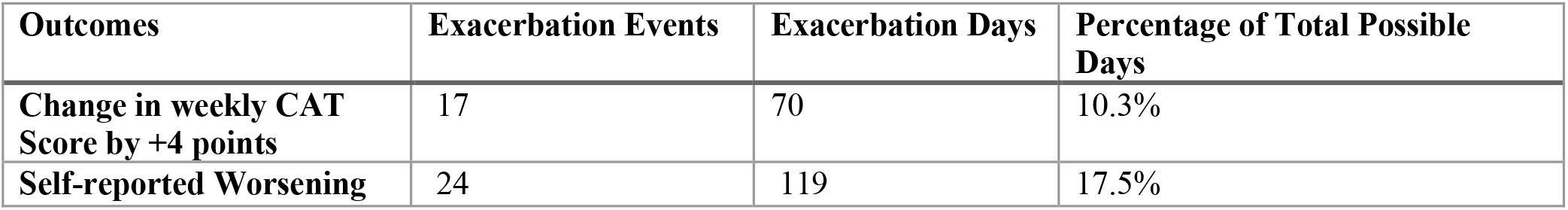
List of Number of Exacerbation Events and Event Days based on Outcome Types.

### 3.2 Prediction Probability

Figure 4 and Figure 5 illustrate examples of an XGBoost model trained on CAT score changes, with the data split and optimized parameters detailed in Table 1. Figure 4 shows a patient in the training set (s18), while Figure 5 shows a patient in the test set (s10).

**Figure 4.**
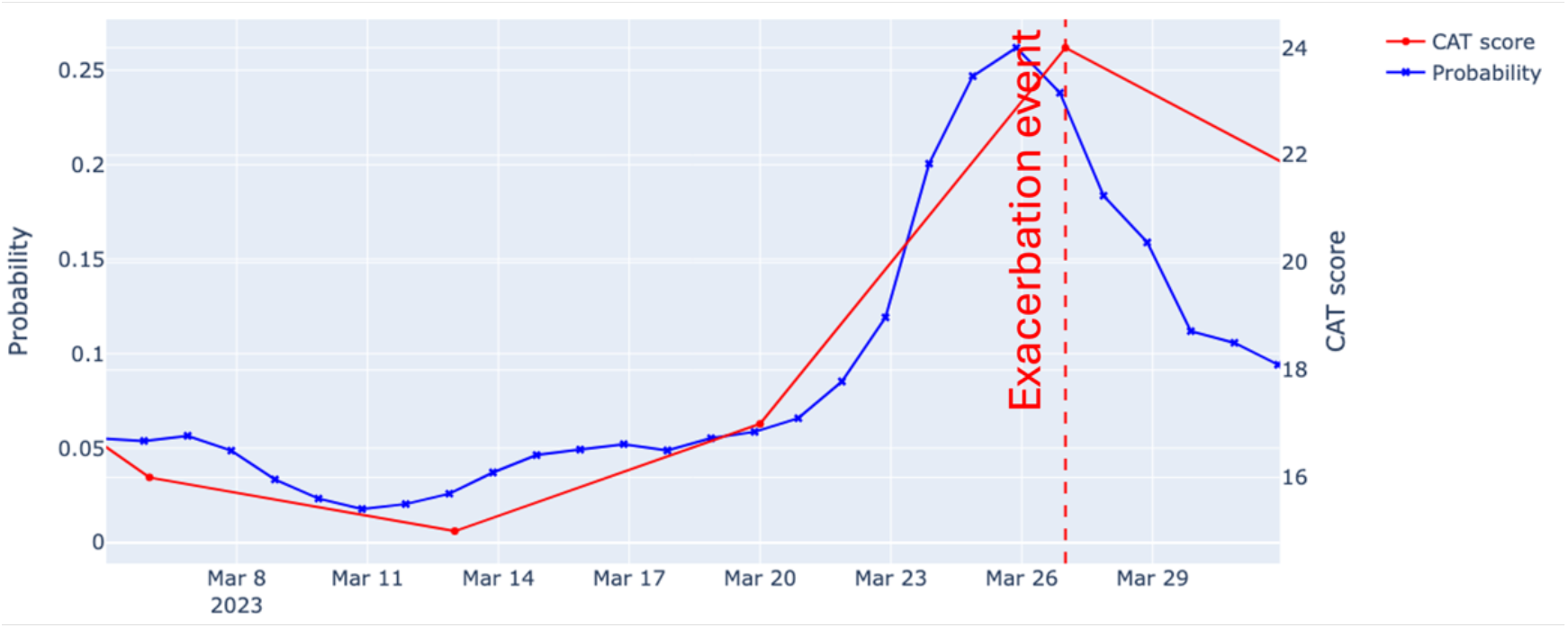
Prediction probability of a patient in the train set – s18.

**Figure 5.**
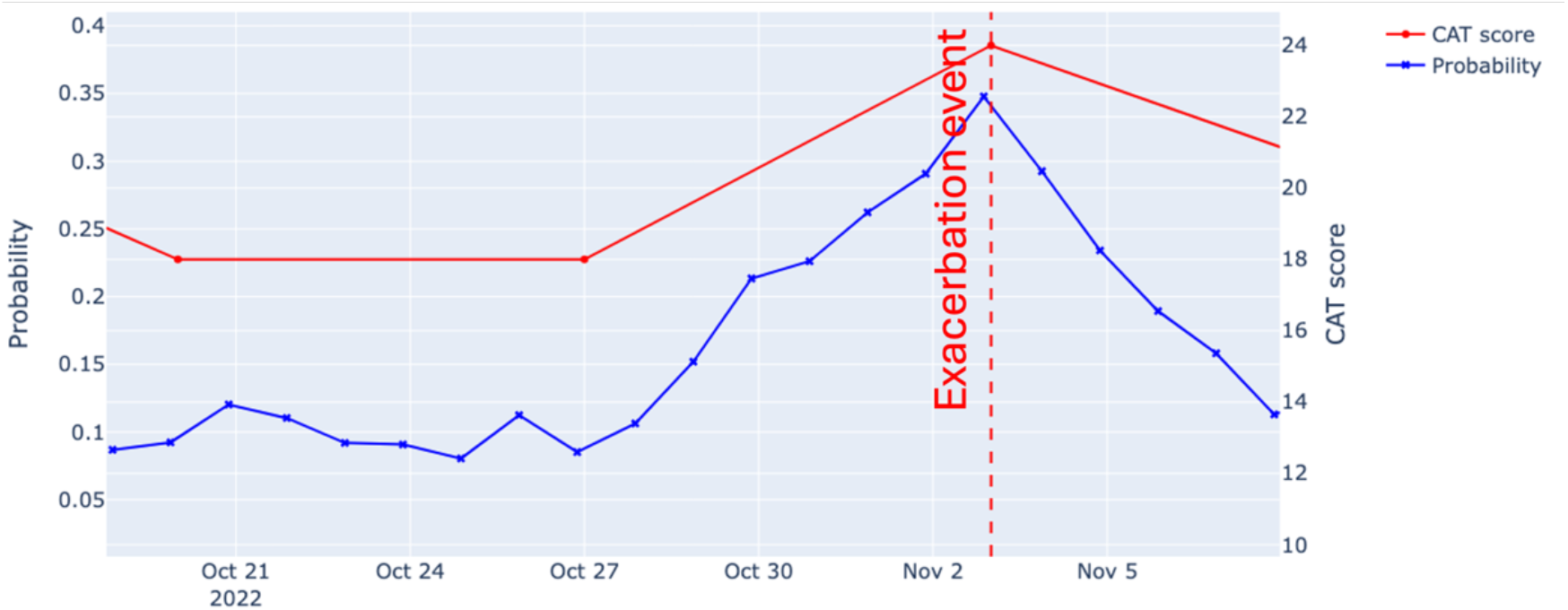
Prediction probability of a patient in the test set – s10

### 3.3 XGBoost vs Logistic Regression

The optimum data cutting parameters were chosen such that an average of AUC ROC (Area Under Curve of Receiver Operating Characteristic) across all the train-test set was maximized. Table 6 and Table 7 show the performance of self-reported and CAT changes outcome respectively using respiratory rate as a predictor.

**Table 6.**
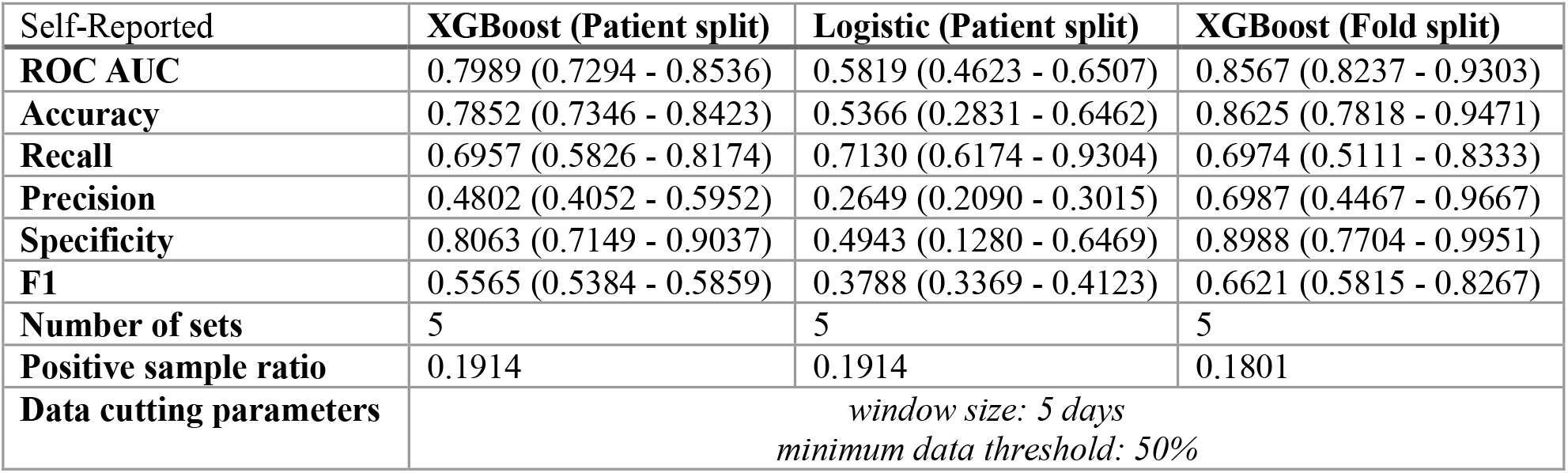
Performance of the Models on Self-reported Outcome.

| Self-Reported | XGBoost (Patient split) | Logistic (Patient split) | XGBoost (Fold split) |
| --- | --- | --- | --- |
| **ROC AUC** | 0.7989 (0.7294 - 0.8536) | 0.5819 (0.4623 - 0.6507) | 0.8567 (0.8237 - 0.9303) |
| **Accuracy** | 0.7852 (0.7346 - 0.8423) | 0.5366 (0.2831 - 0.6462) | 0.8625 (0.7818 - 0.9471) |
| **Recall** | 0.6957 (0.5826 - 0.8174) | 0.7130 (0.6174 - 0.9304) | 0.6974 (0.5111 - 0.8333) |
| **Precision** | 0.4802 (0.4052 - 0.5952) | 0.2649 (0.2090 - 0.3015) | 0.6987 (0.4467 - 0.9667) |
| **Specificity** | 0.8063 (0.7149 - 0.9037) | 0.4943 (0.1280 - 0.6469) | 0.8988 (0.7704 - 0.9951) |
| **F1** | 0.5565 (0.5384 - 0.5859) | 0.3788 (0.3369 - 0.4123) | 0.6621 (0.5815 - 0.8267) |
| **Number of sets** | 5 | 5 | 5 |
| **Positive sample ratio** | 0.1914 | 0.1914 | 0.1801 |
| **Data cutting parameters** | window size: 5 days<br>minimum data threshold: 50% |  |  |

**Table 7.** Performance of the Models on CAT Score Changes Outcome.

| CAT score changes | XGBoost (Patient split) | Logistic (Patient split) | XGBoost (Fold split) |
| --- | --- | --- | --- |
| **ROC AUC** | 0.8720 (0.7204 - 0.9423) | 0.5682 (0.4601 - 0.6631) | 0.8286 (0.7118 - 0.8764) |
| **Accuracy** | 0.8909 (0.6731 - 0.9515) | 0.5607 (0.3177 - 0.9186) | 0.9051 (0.8743 - 0.9344) |
| **Recall** | 0.6667 (0.4615 - 0.9154) | 0.7500 (0.1538 - 1.0000) | 0.5846 (0.3846 - 0.8615) |
| **Precision** | 0.6581 (0.2380 - 1.0000) | 0.3101 (0.1260 - 1.0000) | 0.5740 (0.4429 - 0.7247) |
| **Specificity** | 0.9149 (0.6450 - 1.0000) | 0.5397 (0.2433 - 1.0000) | 0.9412 (0.8757 - 0.9809) |
| **F1** | 0.6032 (0.3761 - 0.7044) | 0.2529 (0.2205 - 0.2881) | 0.5477 (0.4478 - 0.6504) |
| **Number of sets** | 12 | 12 | 5 |
| **Positive sample ratio** | 0.0972 | 0.0972 | 0.1012 |
| **Data cutting parameters** | window size: 4 days<br>minimum data threshold: 25% |  |  |

### 3.4 Respiratory Rate vs Expiratory Time

The performance of the XGBoost models using only respiratory rate and only expiratory time as predictors is compared in Table 8 and Table 9. The expiratory time used as a predictor is the ratio between expiratory time and total breath time. It is important to note that the number of test sets and the positive sample ratio between respiratory rate and expiratory time differ due to varying amounts of missing data in these predictors. Consequently, under the minimum data threshold condition, some vector samples are discarded. Additionally, with the stratified split condition, the test sets and positive sample ratios also vary.

**Table 8.** Performance of the Models on Self-reported Outcome.

| Self-reported | RR (Patient split) | Expiratory Time (Patient split) |
| --- | --- | --- |
| **ROC AUC** | 0.7989 (0.7294 - 0.8536) | 0.6516 (0.6067 - 0.6878) |
| **Accuracy** | 0.7852 (0.7346 - 0.8423) | 0.4182 (0.3437 - 0.5429) |
| **Recall** | 0.6957 (0.5826 - 0.8174) | 0.8627 (0.6294 - 1.0000) |
| **Precision** | 0.4802 (0.4052 - 0.5952) | 0.2393 (0.2328 - 0.2485) |
| **Specificity** | 0.8063 (0.7149 - 0.9037) | 0.3071 (0.1780 - 0.5215) |
| **F1** | 0.5565 (0.5384 - 0.5859) | 0.3697 (0.3535 - 0.3806) |
| **Number of sets** | 5 | 3 |
| **Positive sample ratio** | 0.1914 | 0.2000 |
| **Data cutting parameters** | window size: 5 days<br>minimum data threshold: 50% |  |

**Table 9.**
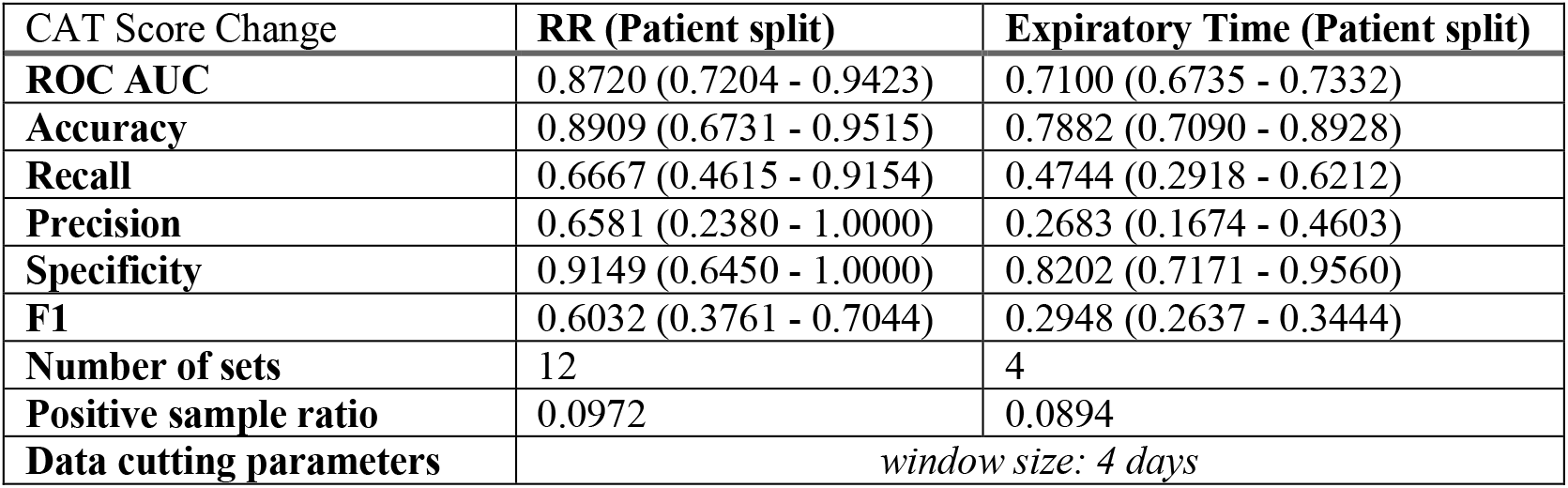
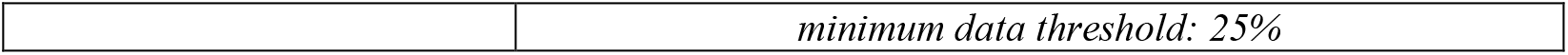
Performance of the Models on CAT Score Changes Outcome.

| CAT Score Change | RR (Patient split) | Expiratory Time (Patient split) |
| --- | --- | --- |
| **ROC AUC** | 0.8720 (0.7204 - 0.9423) | 0.7100 (0.6735 - 0.7332) |
| **Accuracy** | 0.8909 (0.6731 - 0.9515) | 0.7882 (0.7090 - 0.8928) |
| **Recall** | 0.6667 (0.4615 - 0.9154) | 0.4744 (0.2918 - 0.6212) |
| **Precision** | 0.6581 (0.2380 - 1.0000) | 0.2683 (0.1674 - 0.4603) |
| **Specificity** | 0.9149 (0.6450 - 1.0000) | 0.8202 (0.7171 - 0.9560) |
| **F1** | 0.6032 (0.3761 - 0.7044) | 0.2948 (0.2637 - 0.3444) |
| **Number of sets** | 12 | 4 |
| **Positive sample ratio** | 0.0972 | 0.0894 |
| **Data cutting parameters** | window size: 4 days |  |

|  |  |
| --- | --- |
|  | *minimum data threshold: 25%* |

For all the results presented in Tables 6-9, the performance metrics, ROC AUC, accuracy, recall, precision, specificity, and F1 score, were calculated using only the test set. These metrics are reported as average values across all possible train-test sets, with their 95% confidence intervals. As described in the previous section, the fold split consistently results in five sets, while the patient split depends on the stratified condition. Detailed information about the train-test sets can be found in Tables A1-A2 in the supplementary data.

### 3.5 Observation Window Size Optimization

For XGBoost models using respiratory rate as a predictor, a grid search on observation window size was performed to find optimal values. The optimization was performed on patient split only. Figure 6 plots the mean AUC ROC across different window sizes with a fixed optimal minimum data threshold for each outcome. For the self-reported outcome, the minimum data threshold was fixed at 50%; for CAT score changes, it was fixed at 25%. We can generally observe that as the window size increases, the performance reduces for both self-reported outcome and CAT score outcome, possibly because of additional noise as the window size increases.

**Figure 6.**
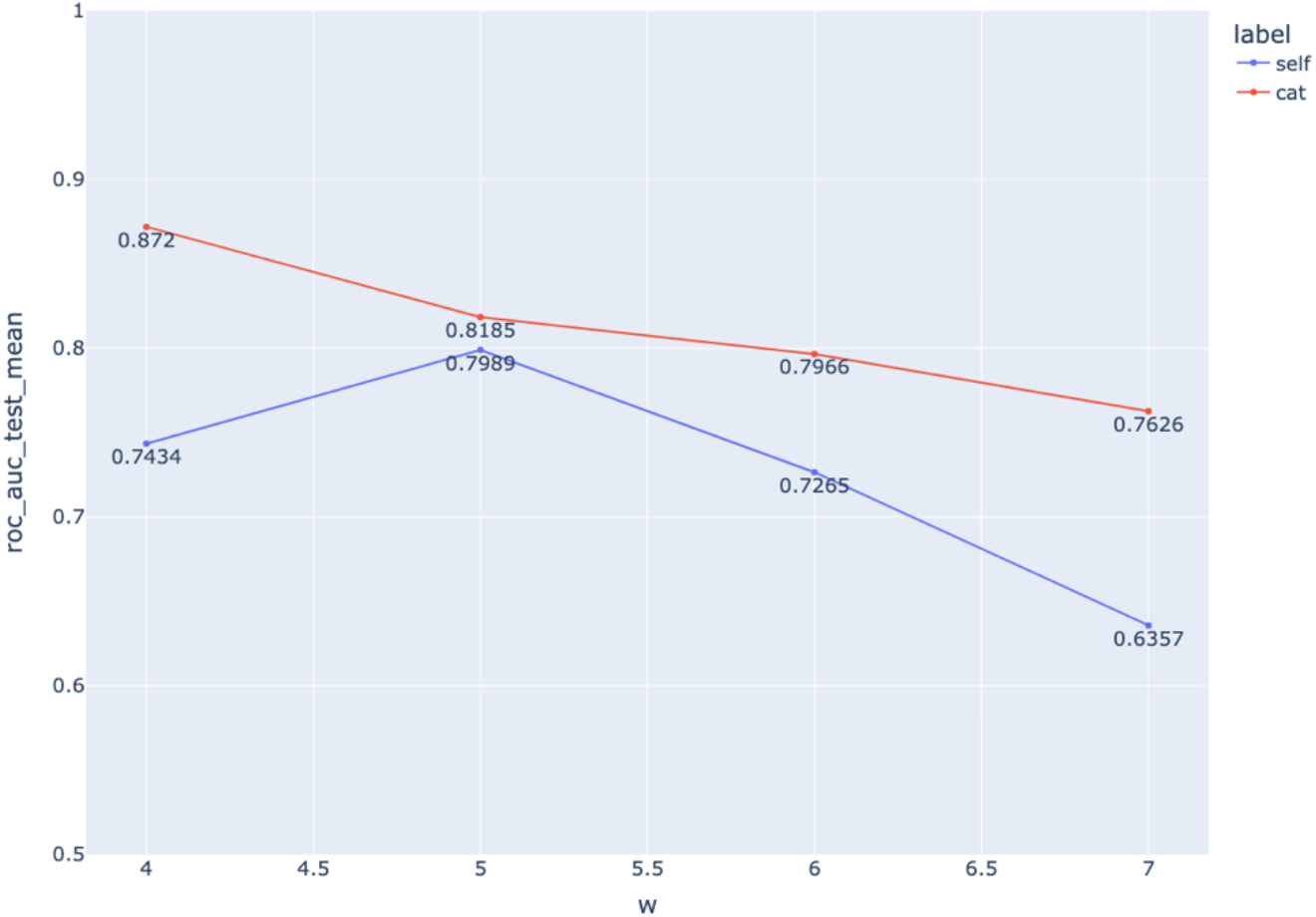
Mean AUC ROC across all test sets with window size of 4, 5, 6 and 7 days

### 3.6 Exacerbation Day Labeling

Although we mainly focus on pre-exacerbation days identification where the five days preceding an exacerbation event and the day of the event itself are labeled as exacerbation days, we performed preceding and succeeding days sweep to study the effect of labeling strategy. Figure 7 shows the average AUC ROC for all combinations of preceding and succeeding days up to 5 days. AUC ROC was calculated based on the optimum set of parameters for each outcome (self-reported: window size = 5 days, minimum data threshold = 50%, CAT score changes: window size = 4 days, minimum data threshold = 25%).

**Figure 7.**
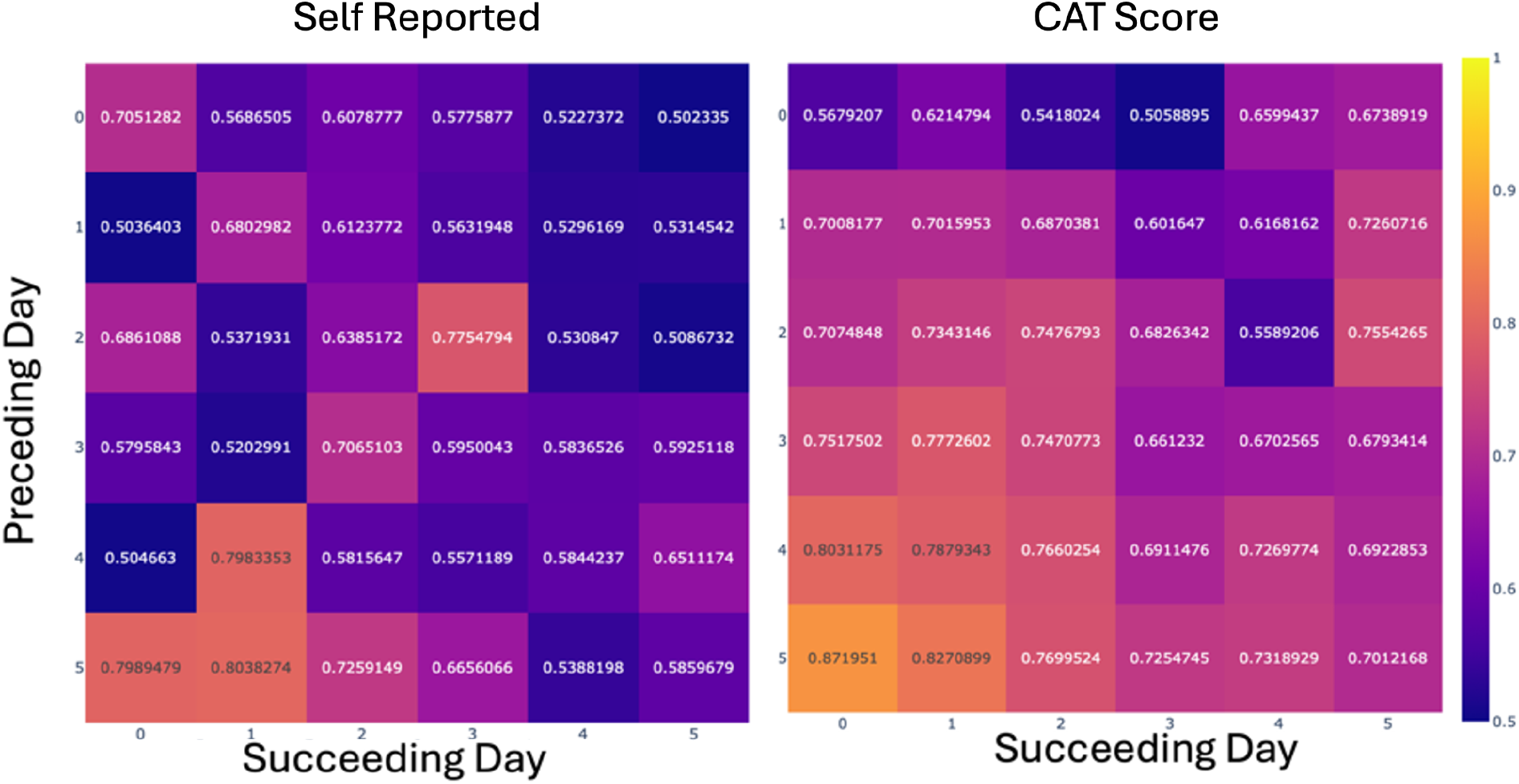
Mean of AUC ROC of all combinations of preceding days and succeeding days.

## 4. Discussion

From Figure 4 and Figure 5, the trend of the predicted probability is similar to that of the CAT scores, indicating that the model implicitly learned the trend of the CAT scores, even though the training labels were binary. The probability starts to increase gradually around 7 days before the exacerbation events which potentially can be used for early identification.

According Table 6 and Table 7, XGBoost achieves an AUC ROC of 0.7989 compared to logistic regression’s 0.5819 for the self-reported outcome. XGBoost also attains AUC ROC scores of 0.8720 for CAT score changes, while logistic regression achieves 0.5682 for the same outcome. XGBoost consistently outperforms logistic regression across all two outcomes. As a tree-based model, XGBoost is likely better able to learn complex patterns in respiratory rates, unlike logistic regression, which is a linear model that might only weigh respiratory rates at each time point. Additionally, XGBoost can naturally handle missing data, whereas logistic regression requires data imputation. It is worth noting that the optimal data cutting parameters vary for each outcome. An observation window size of 4-5 days is optimal, while the optimal minimum data threshold ranges widely from 25% (for CAT score changes) to 50% (for self-reported outcomes).

Table 8 and Table 9 compare the performance of models using respiratory rate and expiratory time as predictors. Respiratory rate outperforms expiratory time, suggesting that exacerbation events in COPD patients have a greater impact on respiratory rate patterns than on expiratory time patterns. However, the AUC ROC scores of 0.6516 for self-reported outcomes and 0.7100 for CAT score changes using expiratory time indicate that the model was able to learn some patterns from this predictor, and hence, this predictor might still be useful when adopted in subsequent multi-parametric models.

For observation window size optimization, Figure 6 shows that an observation window size of five days is optimal for the self-reported outcome, while a window size of four days is optimal for CAT score changes. With this optimal observation window size, we can conclude that for the self-reported outcome, the model needs respiratory rate data for at least five days. For CAT score changes, the model requires at least four days.

Finally, the labeling of preceding and succeeding days of exacerbation events was tested. Figure 7 shows that, in general, a longer duration of preceding days is more effective. For the self-reported outcome, using five preceding days with one succeeding day yields the best results, while for CAT score changes, using five preceding days without any succeeding days performs best, suggesting that in general, identifying pre-exacerbation days is easier than identifying post-exacerbation days. Although this might seem counterintuitive, the impact of interventions following exacerbation events is unknown. It is possible that interventions may alter respiratory rate patterns after exacerbation events, making it more challenging for the model to learn and recognize post-exacerbation patterns. Additionally, longer durations of preceding days result in better performance compared to shorter durations because there are more positive vector samples, which reduces data imbalance. In other words, a larger window gives the model more opportunities to predict correctly.

There are limitations to this study. First, symptom scores were taken on a weekly basis. While this may be sufficient for a preliminary study, subsequent research should account for daily symptom scores to provide more granular data. Second, interventions were not recorded in this study. Although the labeling strategies are retrospective and do not account for days preceding the exacerbations, future studies should incorporate interventions to provide a more comprehensive understanding of their impact on respiratory rate patterns post exacerbations.

## 5. Conclusion

This study demonstrated the feasibility and effectiveness of using aggregated respiratory rate and expiratory time data, combined with machine learning models, to predict pre-exacerbation days in COPD patients. By utilizing XGBoost models, the research highlighted the critical role of optimizing data cutting parameters, such as observation window size and minimum data threshold, which significantly influenced the performance metrics including ROC AUC, accuracy, recall, and precision. The ability to predict days leading up to COPD exacerbations provides a valuable tool for early intervention and proactive patient management.

The focus on predicting pre-exacerbation days is particularly important, as it allows for timely clinical interventions that can prevent exacerbation events. By accurately identifying these critical periods, healthcare providers can take preemptive actions, such as adjusting medication or increasing monitoring, thereby potentially reducing hospital admissions and improving patient outcomes. This proactive approach not only enhances patient quality of life but also reduces healthcare costs associated with emergency treatments and hospitalizations.

Future research should expand on these promising findings by incorporating larger, more diverse datasets and exploring additional physiological parameters to validate and refine the models. Integrating patient-reported outcomes and other health metrics could further enhance the robustness and applicability of the predictive models. Overall, this study marks a significant advancement in leveraging machine learning for improved COPD management, with promising implications for broader applications in respiratory and other chronic diseases, emphasizing the potential for early intervention and better patient care.

## Data Availability

All data produced in the present study are available upon reasonable request to the authors.

## 6. Acknowledgements

This study is funded by the Agency for Science Technology and Research Grant Funding. We would like to acknowledge our collaborators M. Chiu and J. Rooney from Massachusetts General Hospital, A. Attia and G. Balasundaram from A*STAR Skin Research Laboratory, Agency for Science Technology and Research (Singapore), and T. Antoni, M. Belman, C. Cooper, B. Celli from Respiree Pte Ltd. (Singapore).

## Notes

### Competing Interest Statement

The authors have declared no competing interest.

### Author Declarations

We enrolled 17 subjects diagnosed with COPD at the Massachusetts General Hospital (MGH) and Salem Hospital in a prospective cohort/diagnostic accuracy study approved under the Mass General Brigham Institutional Review Board (IRB Protocol #2021P000895, Clinicaltrials.gov ID: NCT04825067).

## References

[1] “National Hospital Ambulatory Medical Care Survey: 2021 Emergency Department Summary Tables,” National Center for Health Statistics, Oct. 2023.

[2] Y. Liu, S. A. Carlson, K. B. Watson, F. Xu, and K. J. Greenlund, “Trends in the Prevalence of Chronic Obstructive Pulmonary Disease Among Adults Aged ≥18 Years – United States, 2011–2021,” MMWR Morb. Mortal. Wkly. Rep., vol. 72, no. 46, pp. 1250–1256, Nov. 2023, doi: 10.15585/mmwr.mm7246a1.

[3] N. Tzanakis Nikos, P. Fotis, and G. Hillas, COPD, vol. Volume 11, pp. 1579–1586, July 2016, doi: 10.2147/COPD.S106160.

[4] H. Whittaker et al., “Frequency and Severity of Exacerbations of COPD Associated with Future Risk of Exacerbations and Mortality: A UK Routine Health Care Data Study,” COPD, vol. Volume 17, pp. 427–437, Mar. 2022, doi: 10.2147/COPD.S346591.

[5] A. J. Mackay et al., “Patient-reported Outcomes for the Detection, Quantification, and Evaluation of Chronic Obstructive Pulmonary Disease Exacerbations,” Am J Respir Crit Care Med, vol. 198, no. 6, pp. 730–738, Sept. 2018, doi: 10.1164/rccm.201712-2482CI.

[6] N. W. Johnston et al., “Detection of COPD Exacerbations and Compliance With Patient-Reported Daily Symptom Diaries Using a Smartphone-Based Information System,” Chest, vol. 144, no. 2, pp. 507–514, Aug. 2013, doi: 10.1378/chest.12-2308.

[7] D. Feliz-Rodriguez et al., “Evolution of the COPD Assessment Test Score during Chronic Obstructive Pulmonary Disease Exacerbations: Determinants and Prognostic Value,” Canadian Respiratory Journal, vol. 20, no. 5, Jan. 2013, doi: 10.1155/2013/398120.

[8] M. G. Crooks et al., “Evidence generation for the clinical impact of myCOPD in patients with mild, moderate and newly diagnosed COPD: a randomised controlled trial,” ERJ Open Res, vol. 6, no. 4, pp. 00460–02020, Oct. 2020, doi: 10.1183/23120541.00460-2020.

[9] J. Alladina et al., “Observational study of the Amaze^TM^ asthma disease management platform,” DIGITAL HEALTH, vol. 10, p. 20552076241282380, Jan. 2024, doi: 10.1177/20552076241282380.

[10] S. A. Shah, C. Velardo, A. Farmer, and L. Tarassenko, “Exacerbations in Chronic Obstructive Pulmonary Disease: Identification and Prediction Using a Digital Health System,” J Med Internet Res, vol. 19, no. 3, p. e69, Mar. 2017, doi: 10.2196/jmir.7207.

[11] A. Al Rajeh and J. Hurst, “Monitoring of Physiological Parameters to Predict Exacerbations of Chronic Obstructive Pulmonary Disease (COPD): A Systematic Review,” JCM, vol. 5, no. 12, p. 108, Nov. 2016, doi: 10.3390/jcm5120108.

[12] R. Rodriguez-Roisin et al., “Daily home-based spirometry during withdrawal of inhaled corticosteroid in severe to very severe chronic obstructive pulmonary disease,” Int J Chron Obstruct Pulmon Dis, vol. 11, pp. 1973–1981, 2016, doi: 10.2147/COPD.S106142.

[13] P. L. Enright, M. Studnicka, and J. Zielinski, “Spirometry to detect and manage chronic obstructive pulmonary disease and asthma in the primary care setting,” in Lung Function Testing, 1st ed., R. Gosselink and H. Stam, Eds., European Respiratory Society, 2005. doi: 10.1183/1025448x.00031001.

[14] N. Sabati, M. Snyder, C. Edin-Stibbe, B. Lindgren, and S. Finkelstein, “Facilitators and barriers to adherence with home monitoring using electronic spirometry,” AACN Clin Issues, vol. 12, no. 2, pp. 178–185, May 2001, doi: 10.1097/00044067-200105000-00002.

[15] Y. H. Khor et al., “Assessment of Home-based Monitoring in Adults with Chronic Lung Disease: An Official American Thoracic Society Research Statement,” Am J Respir Crit Care Med, vol. 211, no. 2, pp. 174–193, Feb. 2025, doi: 10.1164/rccm.202410-2080ST.

[16] A. M. Yañez et al., “Monitoring Breathing Rate at Home Allows Early Identification of COPD Exacerbations,” Chest, vol. 142, no. 6, pp. 1524–1529, Dec. 2012, doi: 10.1378/chest.11-2728.

[17] S. Takahashi, E. Nakazawa, S. Ichinohe, A. Akabayashi, and A. Akabayashi, “Wearable Technology for Monitoring Respiratory Rate and SpO2 of COVID-19 Patients: A Systematic Review,” Diagnostics (Basel), vol. 12, no. 10, p. 2563, Oct. 2022, doi: 10.3390/diagnostics12102563.

[18] “Global Strategy for the Diagnosis, Management, and Prevention of Chronic Obstructive Pulmonary Disease,” Global Initiative for Chronic Obstructive Lung Disease, 2025.

[19] G. Singh, A. Tee, T. Trakoolwilaiwan, A. Taha, and M. Olivo, “Method of respiratory rate measurement using a unique wearable platform and an adaptive optical-based approach,” ICMx, vol. 8, no. 1, p. 15, Dec. 2020, doi: 10.1186/s40635-020-00302-6.

[20] P. A. Harris, R. Taylor, R. Thielke, J. Payne, N. Gonzalez, and J. G. Conde, “Research electronic data capture (REDCap)--a metadata-driven methodology and workflow process for providing translational research informatics support,” J Biomed Inform, vol. 42, no. 2, pp. 377–381, Apr. 2009, doi: 10.1016/j.jbi.2008.08.010.

[21] P. A. Harris et al., “The REDCap consortium: Building an international community of software platform partners,” Journal of Biomedical Informatics, vol. 95, p. 103208, July 2019, doi: 10.1016/j.jbi.2019.103208.

[22] S. S. C. Kon et al., “Minimum clinically important difference for the COPD Assessment Test: a prospective analysis,” The Lancet Respiratory Medicine, vol. 2, no. 3, pp. 195–203, Mar. 2014, doi: 10.1016/S2213-2600(14)70001-3.

[23] F.-A. Coutu, O. C. Iorio, and B. A. Ross, “Remote patient monitoring strategies and wearable technology in chronic obstructive pulmonary disease,” Front. Med., vol. 10, p. 1236598, Aug. 2023, doi: 10.3389/fmed.2023.1236598.

[24] “EP12 | Evaluation of Qualitative, Binary Output Examination Performance.” Accessed: Oct. 22, 2025. [Online]. Available: https://clsi.org/shop/standards/ep12/

