## Supplementary data for "Using Hourly Aggregated Respiratory Rate and Expiratory Time with Machine Learning to Identify Remote COPD Exacerbations"

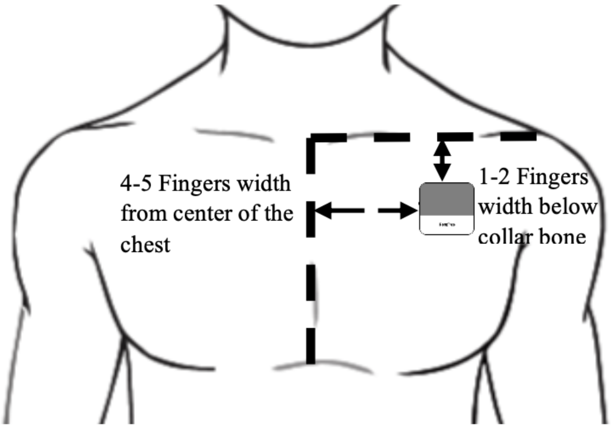


**Figure A.** Optimal placement of the wearable device for monitoring.

| Train set | Number of vector samples | Positive samples (%) | Test set | Number of vector samples | Positive samples (%) |
|---|---|---|---|---|---|
| **Patient-Fold Split** | | | | | |
| ('s10', 's13', 's16', 's20', 's21', 's22', 's26', 's27', 's29', 's36', 's37', 's39') | 375 | 17.600 | ('s05', 's18', 's28') | 119 | 19.328 |
| ('s10', 's13', 's16', 's20', 's21', 's22', 's28', 's29', 's36', 's37', 's39') | 374 | 17.647 | ('s05', 's18', 's26', 's27') | 120 | 19.167 |
| ('s10', 's13', 's16', 's20', 's21', 's22', 's26', 's28', 's36', 's37', 's39') | 372 | 17.742 | ('s05', 's18', 's27', 's29') | 122 | 18.852 |
| ('s10', 's13', 's16', 's20', 's21', 's22', 's26', 's28', 's29', 's36', 's39') | 374 | 17.647 | ('s05', 's18', 's27', 's37') | 120 | 19.167 |
| ('s10', 's13', 's16', 's20', 's21', 's22', 's26', 's28', | 374 | 17.647 | ('s05', 's18', 's27', 's39') | 120 | 19.167 |

| | | | | | |
|---|---|---|---|---|---|
| 's29', 's36', 's37') | | | | | |
| **5-Fold Split** | | | | | |
| **Fold 1** | 395 | 17.975 | | 99 | 18.182 |
| **Fold 2** | 395 | 17.975 | | 99 | 18.182 |
| **Fold 3** | 395 | 17.975 | | 99 | 18.182 |
| **Fold 4** | 395 | 17.975 | | 99 | 18.182 |
| **Fold 5** | 396 | 18.182 | | 98 | 17.347 |

*Table A1 Patient Split and Fold Split of Self-reported Outcome*

| **Train set** | **Number of vector samples** | **Positive samples (%)** | **Test set** | **Number of vector samples** | **Positive samples (%)** |
|---|---|---|---|---|---|
| **Patient Split** | | | | | |
| ('s13', 's16', 's18', 's20', 's21', 's22', 's26', 's27', 's28', 's29', 's36', 's37', 's39') | 505 | 10.297 | ('s05', 's10') | 137 | 9.489 |
| ('s05', 's13', 's16', 's18', 's21', 's22', 's27', 's28', 's29', 's36', 's37', 's39') | 505 | 10.297 | ('s10', 's20', 's26') | 137 | 9.489 |
| ('s05', 's13', 's16', 's18', 's21', 's22', 's26', 's27', 's29', 's36', 's37', 's39') | 509 | 10.216 | ('s10', 's20', 's28') | 133 | 9.774 |
| ('s05', 's13', 's16', 's18', 's21', 's22', 's26', 's27', 's28', 's29', 's36', 's39') | 512 | 10.156 | ('s10', 's20', 's37') | 130 | 10.000 |
| ('s05', 's13', 's16', 's18', 's21', 's22', 's26', 's28', 's29', 's37', 's39') | 512 | 10.156 | ('s10', 's20', 's27', 's36') | 130 | 10.000 |

| | | | | | |
|---|---|---|---|---|---|
| ('s05', 's13', 's16', 's18', 's21', 's22', 's26', 's28', 's29', 's36', 's37') | 503 | 10.338 | ('s10', 's20', 's27', 's39') | 139 | 9.353 |
| ('s05', 's13', 's16', 's18', 's21', 's22', 's26', 's27', 's29', 's37', 's39') | 504 | 10.317 | ('s10', 's20', 's28', 's36') | 138 | 9.420 |
| ('s05', 's13', 's16', 's18', 's21', 's22', 's26', 's27', 's28', 's29', 's39') | 507 | 10.256 | ('s10', 's20', 's36', 's37') | 135 | 9.630 |
| ('s05', 's13', 's16', 's18', 's21', 's22', 's26', 's27', 's28', 's29', 's37') | 511 | 10.176 | ('s10', 's20', 's36', 's39') | 131 | 9.924 |
| ('s05', 's13', 's16', 's18', 's20', 's21', 's22', 's29', 's36', 's37', 's39') | 512 | 10.156 | ('s10', 's26', 's27', 's28') | 130 | 10.000 |
| ('s05', 's13', 's16', 's18', 's20', 's21', 's22', 's27', 's29', 's36', 's39') | 507 | 10.256 | ('s10', 's26', 's28', 's37') | 135 | 9.630 |
| ('s05', 's13', 's16', 's18', 's20', 's21', 's22', 's27', 's29', 's36', 's37') | 511 | 10.176 | ('s10', 's26', 's28', 's39') | 131 | 9.924 |
| **Fold Split** | | | | | |
| **Fold 1** | 513 | 10.136 | | 129 | 10.078 |
| **Fold 2** | 513 | 10.136 | | 129 | 10.078 |
| **Fold 3** | 514 | 10.117 | | 128 | 10.156 |
| **Fold 4** | 514 | 10.117 | | 128 | 10.156 |

| | | | | | |
|---|---|---|---|---|---|
| **Fold 5** | 514 | 10.117 | | 128 | 10.156 |

*Table A2 Patient Split and Fold Split of CAT Score Changes Outcome*

### Equations

$TPR = \frac{TP}{TP + FN}$ (E1) and $FPR = \frac{FP}{TN + FP}$ (E2), where *TP* is the number of true positives, *TN* true negatives, *FP* false positives and *FN* false negatives.

$Accuracy = \frac{TP + TN}{TP + TN + FP + FN}$ (E3)

$Recall = TPR = \frac{TP}{TP + FN}$ (E4)

$Precision = \frac{TP}{TP + FP}$ (E5)

$Specificity = \frac{TN}{TN + FP}$ (E6)

$F1 = \frac{2 \times Precision \times Recall}{Precision + Recall}$ (E7)
